# OCCULT INTRACRANIAL INJURIES ON COMPUTED TOMOGRAPHY (CT) HEAD SCANS IN INFANTS INVESTIGATED FOR SUSPECTED PHYSICAL ABUSE: A RETROSPECTIVE REVIEW

**DOI:** 10.1101/2020.05.21.20106948

**Authors:** H Daley, H Smith, S McEvedy, R King, ET Andrews, F Hawkins, N Guppy, T Kiryazova, R Macleod, E Blake, R Harrison

## Abstract

**Background:** United Kingdom national guidelines recommend that investigation of infants (aged <12 months) with suspected physical abuse should always include computed tomography (CT) head scans. Studies report a range of yields for occult intracranial injuries in infants.

**Aims:** To gauge the yield of occult intracranial injuries on CT head scans in infants who underwent radiological investigations for suspected physical abuse, and compare selected demographic, clinical and radiological features in infants with and without intracranial injuries.

**Method:** A retrospective cross-sectional review of infants investigated for suspected physical abuse in Wessex, England. The main outcome measure was yield of occult intracranial injuries on CT head scan. Occult injuries were defined as previously unsuspected CT head scan findings of postnatal intracranial injury.

**Results:** Of 363 CT head scans meeting study criteria, 68 were in infants with neurological signs or skull fractures, 36 of these had intracranial injuries. Of the 295 without neurological signs or skull fractures, just 1 infant had CT features of an intracranial injury. This was the only occult intracranial injury found. It was a small cortical haemorrhage found to be consistent with a contracoup injury from an accidental fall. No additional demographic, clinical or radiological features were associated with intracranial injury.

**Conclusion:** In suspected physical abuse, CT head scans should be carried out in infants who present with neurological signs, or who have skull fractures identified on X-ray. However, we question the benefit of performing CT head scans routinely in infants who have no signs of head injury.

## INTRODUCTION

In the United Kingdom (U.K.), national guidance on radiological investigation for suspected physical abuse in children recommends that imaging ‘should always include…computed tomography (CT) head scan in children under one year old’. [1, 2] Abusive head trauma is the leading cause of death from physical abuse in this age group. [3]

CT head scans have been preferred over magnetic resonance imaging (MRI) for initial assessment of physical abuse because sedation is usually not required and because of availability, shorter scan times, and better detection of bone trauma and recent bleeding. Risks of radiation must be taken into account when requesting CT scans in infants (age <12 months) because, although risks are small, exposure to radiation is known to increase the risk of cancer, particularly in children.[4. 5] The precise level of risk in infants is difficult to determine, though they are likely to have higher levels of risk than other ages.[6]

Studies reporting frequency of occult head injuries in infants with suspected physical abuse differ in their definition of occult and in populations studied, with reported yields ranging from 4% to 38%. [7-12] Information about yield is important in balancing risks and benefits of medical investigations. The Royal College of Paediatrics and Child Health calls for ‘further research. to determine the positive diagnostic yield of head CT in children less than one year of age’.[3] This study contributes to the body of research aiming to answer this question.

The aims of this study were to gauge the yield of occult intracranial injuries on CT head scans in infants who underwent radiological investigations for suspected physical abuse, and to compare selected demographic, clinical and radiological features in infants with and without intracranial injuries.

## METHODS

A retrospective cross-sectional design was adopted, involving review of records of all infants (children aged from birth until the day before first birthday) who had radiological investigations as part of the assessment for possible physical abuse in a 6-year period from beginning of January 2013 to end of December 2018. This period of time was chosen to maximise the number of study participants and the quality of information held on electronic records.

Study participants included infants referred for child protection medical assessments by social care, and those who presented directly to community or hospital paediatricians in emergency departments, inpatient or outpatient settings. Skeletal surveys and CT head scans were undertaken at the discretion of the clinical teams, taking into account national guidance that, in suspected physical abuse of infants, skeletal survey and CT head scan should be carried out.[2, 13]

Cases were identified at five National Health Service (NHS) hospitals and one NHS community trust in the Wessex region, England. One of the hospitals contains the regional paediatric intensive care unit. Together, the Trusts cover a population of 2.2 million and have approximately 24 000 births/year. [14] In all settings, radiological investigations are requested only after thorough assessment by a senior paediatrician, with skeletal surveys and CT head scans usually requested simultaneously, though they may be carried out in any order. CT head scans are reported at the hospital radiology departments, either by a specialist tertiary paediatric neuro-radiologist or by two subspecialist paediatric radiologists who have access to the specialist paediatric neuro-radiologist when imaging uncertainties arise.

Hospital radiology electronic systems were used to identify infants who had skeletal surveys. A hand search was made to identify those who had skeletal surveys requested for suspected physical abuse. All infants who had CT head scan in addition to skeletal survey were identified for inclusion in the study. A standard, anonymised template was used to extract information from radiology reports and case records, including signs of head injury and the results of radiological investigations. Information was also extracted to identify possible factors associated with abusive head trauma suggested by Rubin et al,[7] including age, facial injuries, occult rib fractures and multiple occult fractures.

Signs of head injury presenting prior to radiological investigations being requested were extracted and described under two main groups: a) neurological signs; and, b) skull fractures. Where both skeletal surveys and CT head scans were requested, skull fractures seen on X-ray were counted as signs of head injury, even if in practice the CT head scan was carried out first. Signs of head injury were assigned in list order, as described in Figure 1, so that infants were counted only once. For the purposes of this study, vomiting, sleepiness without mention of reduced conscious level, excessive crying and irritability were not counted as clinical signs of head injury, as these findings can be attributed to other causes, and may not always lead to consideration of CT head scan.

**Figure 1.**
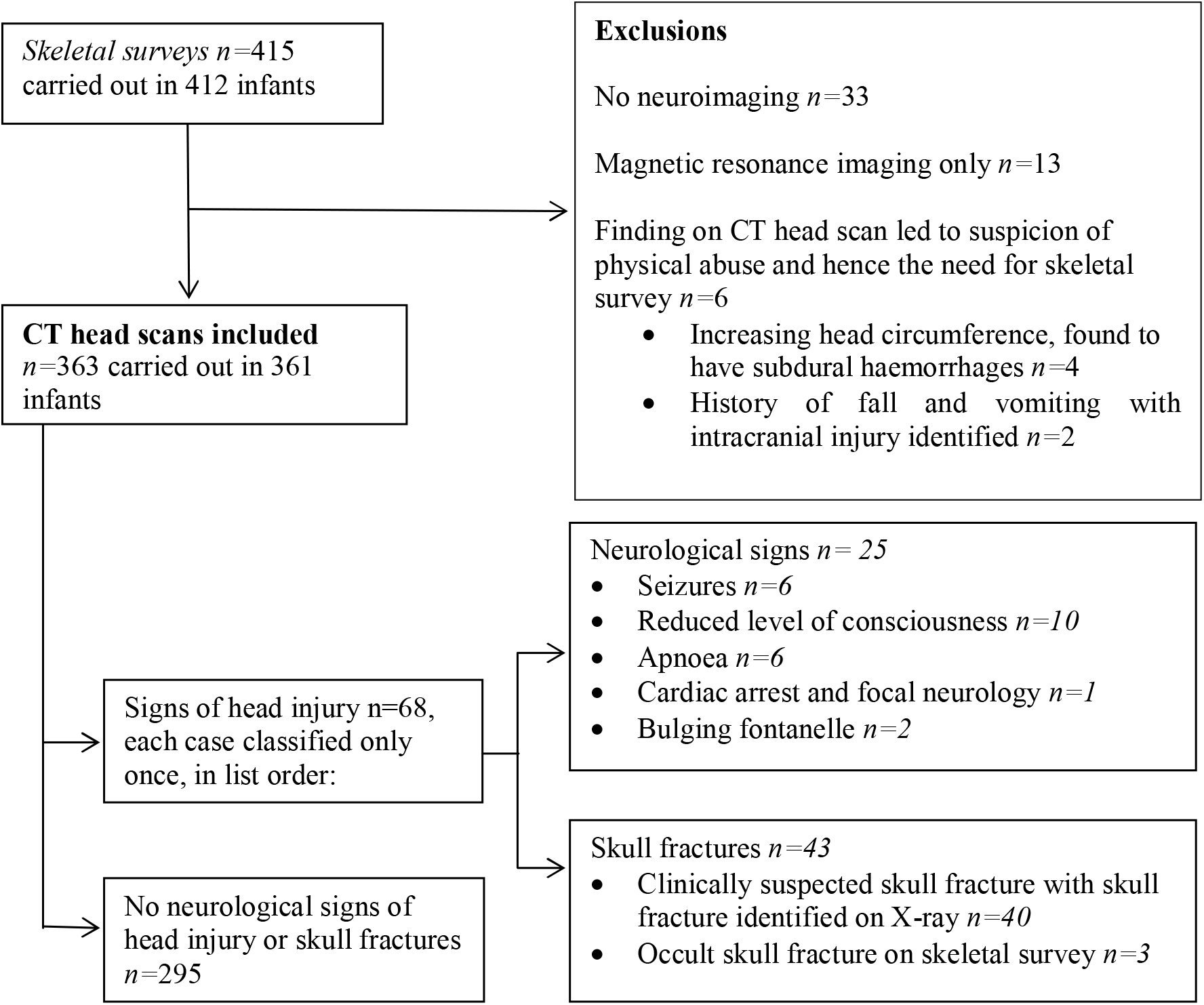
Study flow diagram

The lead author reviewed all clinical and radiological findings. Any ambiguities in interpretation were resolved by discussion with the researchers, who were paediatricians working in each NHS Trust base, and with another researcher independent of the base.

Occult injury was defined as a previously unsuspected CT head scan finding of postnatal intracranial injury. CT scans were considered negative if they were normal or showed only incidental findings that were not suspicious for intracranial injury. Inclusion and exclusion criteria are described in Table 1.

**Table 1.**
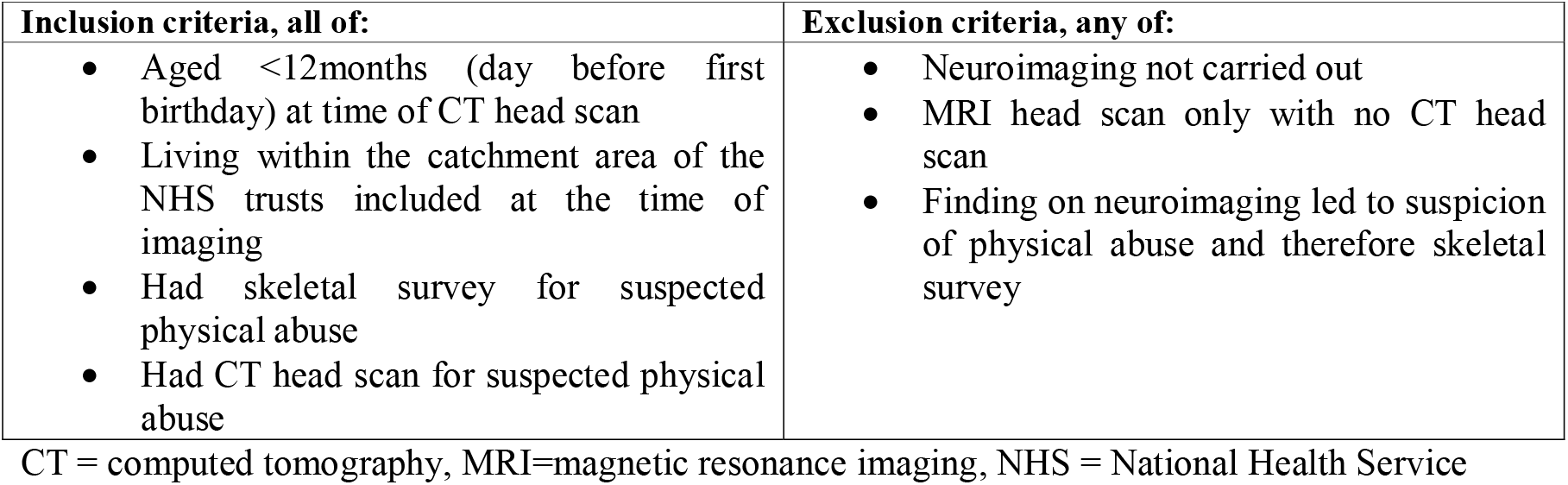
Inclusion and exclusion criteria

### Data analysis

They key outcome measure for this study was the presence of intracranial injury on CT head scan. Potential risk factors for intracranial injury were considered, including demographic, clinical and radiological variables. Data analyses were conducted using IBM SPSS Statistics Version 26. Comparisons were made using chi-square *(X^2^)* or Fishers exact tests for categorical variables.

### Ethical approval

Ethical approval was granted by the NHS Health Research Authority, East of England - Cambridgeshire and Hertfordshire Research Ethics Committee (19/EE/0355).

## RESULTS

Figure 1 shows exclusions from the study and classifications by clinical sign. None of the excluded patients had occult fractures on skeletal survey, and none of the MRI head scans showed occult intracranial injuries.

In total, *n* = 363 CT head scans were included (mean age 3.4, *SD* 2.9 months). Two infants had two separate presentations with CT head scans carried out each time. Neither of these had neurological signs of head injury, skull fractures or intracranial findings on CT head scans.

The characteristics of the study cohort and comparisons between those identified with and without intracranial injuries are shown in Table 2.

**Table 2.**
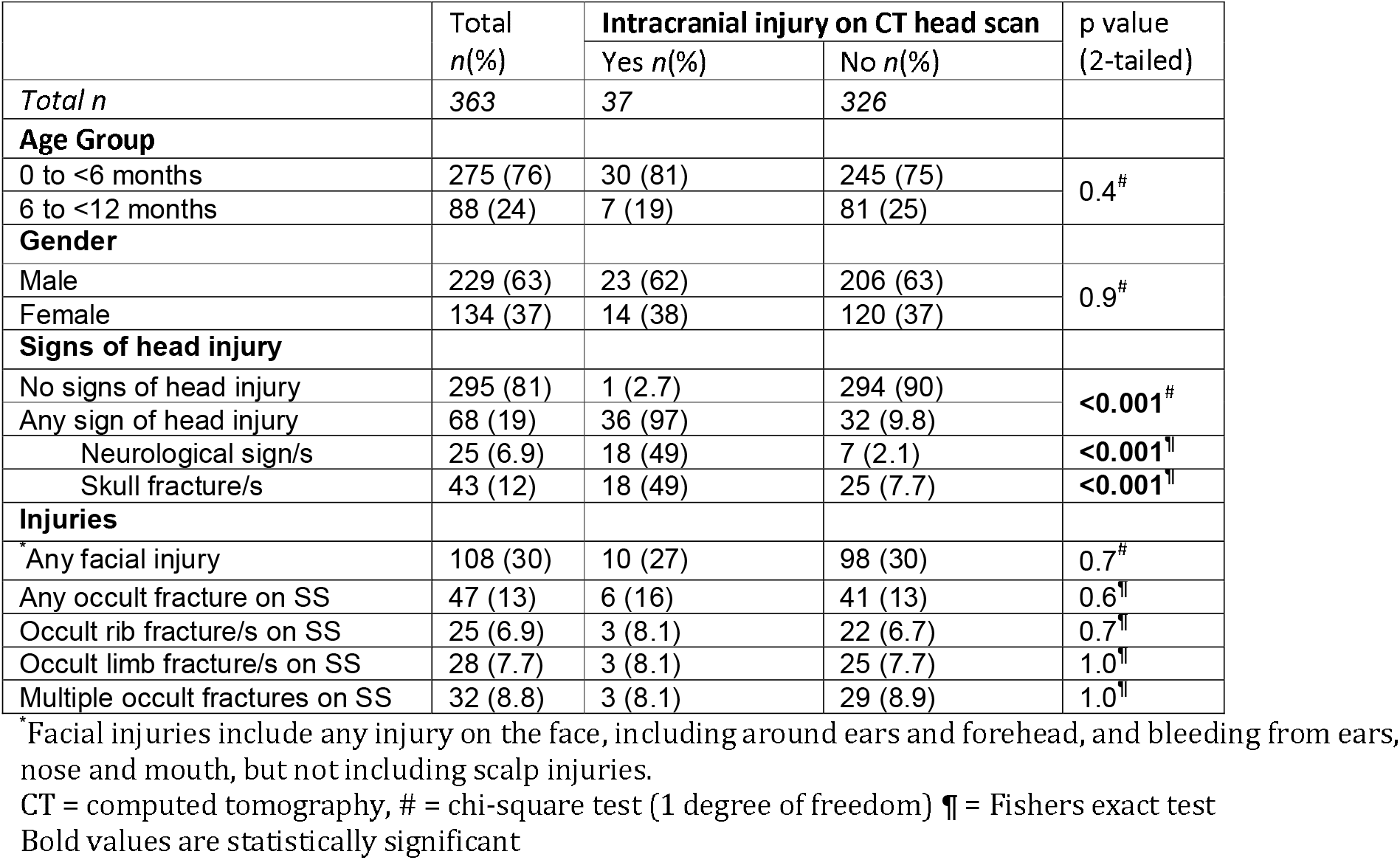
Sample profile and comparison of demographic, clinical and radiological features in infants with and without intracranial injuries on CT head scan

|  | Total<br>*n*(%) | Intracranial injury on CT head scan |  | p value<br>(2-tailed) |
| --- | --- | --- | --- | --- |
|  |  | Yes *n*(%) | No *n*(%) |  |
| *Total n* | 363 | 37 | 326 |  |
| **Age Group** |  |  |  |  |
| 0 to <6 months | 275 (76) | 30 (81) | 245 (75) | 0.4<sup>#</sup> |
| 6 to <12 months | 88 (24) | 7 (19) | 81 (25) |  |
| **Gender** |  |  |  |  |
| Male | 229 (63) | 23 (62) | 206 (63) | 0.9<sup>#</sup> |
| Female | 134 (37) | 14 (38) | 120 (37) |  |
| **Signs of head injury** |  |  |  |  |
| No signs of head injury | 295 (81) | 1 (2.7) | 294 (90) | **<0.001<sup>#</sup>** |
| Any sign of head injury | 68 (19) | 36 (97) | 32 (9.8) |  |
| Neurological sign/s | 25 (6.9) | 18 (49) | 7 (2.1) | **<0.001<sup>¶</sup>** |
| Skull fracture/s | 43 (12) | 18 (49) | 25 (7.7) | **<0.001<sup>¶</sup>** |
| **Injuries** |  |  |  |  |
| Any facial injury | 108 (30) | 10 (27) | 98 (30) | 0.7<sup>#</sup> |
| Any occult fracture on SS | 47 (13) | 6 (16) | 41 (13) | 0.6<sup>¶</sup> |
| Occult rib fracture/s on SS | 25 (6.9) | 3 (8.1) | 22 (6.7) | 0.7<sup>¶</sup> |
| Occult limb fracture/s on SS | 28 (7.7) | 3 (8.1) | 25 (7.7) | 1.0<sup>¶</sup> |
| Multiple occult fractures on SS | 32 (8.8) | 3 (8.1) | 29 (8.9) | 1.0<sup>¶</sup> |

\*Facial injuries include any injury on the face, including around ears and forehead, and bleeding from ears, nose and mouth, but not including scalp injuries.
CT = computed tomography, # = chi-square test (1 degree of freedom) ¶ = Fishers exact test
Bold values are statistically significant

Overall yield of intracranial injury on CT head scan was 10% (95%CI 7-13%). Yield was higher amongst those who had signs of head injury than those who did not (53% vs 0.3%, *X^2^ =* 167.0, *df* = 1, p<0.001). The one occult intracranial injury identified was in an infant aged <6 months, and was reported to be a tiny cortical haemorrhage on a single gyrus which fitted the description of an accidental fall causing contrecoup haemorrhage. The infant had a scalp injury with no skull fracture, and skeletal survey was normal. Investigation was carried out by the multiagency team, and the account of an accidental fall was accepted.

One infant, aged 6 to <12m, presented with limb fractures, and had a linear left occipital bone fracture identified on CT head scan that was seen on skeletal survey only in retrospect. This infant was not found to have intracranial injuries.

Of the 36 infants who presented with intracranial injuries associated with signs of head injury, 18 presented with neurological signs and 19 with skull fractures (one child had both, but the main presentation was with loss of consciousness and they are counted with neurological signs in table 2). Of the 19 with skull fractures, 13 had intracranial injuries directly adjacent to the fracture site only, whilst 6 had more widespread findings of intracranial injury.

## DISCUSSION

This study found that just over half the infants presenting with either neurological signs or skull fractures seen on X-ray had intracranial injuries identified on head CT scan. However, out of 363 CT head scans included in this study, only one occult intracranial injury was identified. Interagency assessment concluded that this injury had been caused by an accidental fall. Therefore, no patients were identified with occult intracranial injuries suggestive of abusive head trauma. This finding is substantially lower than the occult head injury yield reported from other studies.

The yield for occult injuries on skeletal surveys in our study is at the lower end of the range reported, but is in keeping with other published findings,[15-17] suggesting that the population studied is not unusual for infants currently investigated for suspected physical abuse.

U.K. radiology guidelines for the investigation of physical abuse in children, most recently updated in 2018, [2] are based on studies from United States hospitals that report yields ranging from 19-37% for occult injuries on CT head scan. [7-9, 18] In comparison to our study, these studies use different definitions of occult head injury, and there may also be differences in the level of risk for abusive head trauma in included patients.

In determining occult injuries, our study classified patients with any skull fracture seen on X-ray as having a sign of head injury, regardless of whether the CT head scan or skeletal survey were carried out first. However, other published studies[7-12] variably include skull fractures as occult head injuries, and If an intracranial injury is associated with the skull fracture, this would be counted as an occult intracranial injury. In addition, our study did not attempt to ascertain extracranial injuries, as their clinical and forensic value can be difficult to determine. Two of the studies, Rubin et al[7] and Harper et al, [9] include isolated, clinically unsuspected scalp soft tissue swellings as occult signs of head injury. Rubin et al pointed out that although they found occult head injuries in 37.5% of 51 <24month olds meeting high-risk criteria, only five subjects would have missed detection if a skeletal survey had been performed alone. Three of these subjects were described as having isolated scalp swellings.

Different study designs do not allow direct comparison, but the published studies report a wide range for rates of neuroimaging in patients suspected of having physical abuse (36-91%),[7-12] and bias might be expected if higher risk patients were more likely to receive neuroimaging. In addition, patients included in previous studies may have been from a higher risk group than those we currently investigate in the U.K. For example, since National Institute for Health and Care Guidelines were published in 2009,[19] it has become increasingly accepted practice for non-mobile infants with isolated bruises to undergo assessment and radiological investigations for suspected physical abuse.

Since U.K. guidelines were formulated, Shaikh et al[11] reported findings that are in concordance with data from this study. Their yield for occult findings on CT head scan was 7 out of 132 (5%) children under the age of two. Five of the occult findings were skull fractures, one was an orbital fracture, and one was a finding of a right parietal lucency not seen on subsequent MRI scan.

Our study was unable to identify any additional risk factors for intracranial injury apart from neurological signs or skull fractures. These findings are consistent with those reported by Rubin et al[7] and Boehnke et al,[10] which also found no significant differences in presence of facial injuries or skeletal survey findings in children with and without occult head injuries on CT head scan.

### Strengths and Limitations

A strength of this study is that it includes patients from across a range of settings assessed for suspected physical abuse within a population, which helps in generalising the results. Because of the U.K’s universal healthcare system provided by the NHS, we are confident that the sample of infants included in the study is comprehensive. It is unlikely that a significant number of patients would have attended other settings for evaluation of physical abuse, and that any patients who did so would bias the findings of this study. Robust arrangements for radiology reporting are in place within each hospital with quality control for accurate reporting, and there were no observable discrepancies in clinical or radiological findings between the NHS Trusts involved.

Nevertheless, this study does have several limitations. The retrospective study design limits the completeness of the data, and there are risk factors for intracranial injuries which were not collected. For example, data around socio-economic status, and other social risk factors were not included in the data set. Future research could benefit from gathering data prospectively at the time of CT head scan for a wider range of risk factors or by adopting methods which combine data from a range of sources.

The study did not include infants with significant burn injuries, who are admitted to a regional burns centre. Therefore, our findings cannot be generalised to these patients.

There are an unknown number of infants where physical abuse might have been suspected but who had no skeletal survey carried out, and therefore did not meet the study’s inclusion criteria. We cannot exclude the possibility that CT head scan might have picked up occult intracranial injuries in these patients or in those who didn’t receive CT head scans, and so were excluded from the study.

It is possible that a larger sample size in our study would have picked up more occult intracranial injuries. It would remain important for the yield to be taken into account when considering the risks and benefits of CT head scans in infants investigated for suspected physical abuse.

## CONCLUSION

In suspected physical abuse, CT head scans should be carried out in infants who present with neurological signs, or who have skull fractures identified on X-ray. However, we question the benefit of performing CT head scans routinely in infants who have no signs of head injury.

## Data Availability

All data relevant to the study are included in the article

## ACKNOWLEDGEMENTS

We thank Dr Michael Roe, Consultant Paediatrician, Southampton University Hospital Trust for his invaluable help and support with the design and critical review of this study.

